# impaired bed mobility in prediagnostic and de novo Parkinson’s disease

**DOI:** 10.1101/2022.03.08.22272005

**Authors:** Femke Dijkstra, Ilse de Volder, Mineke Viaene, Patrick Cras, David Crosiers

## Abstract

**Background:** Wearable technology research suggests that nocturnal movements are disturbed in early Parkinson’s disease (PD). In this study, we investigate if patients also already experience impaired bed mobility before PD diagnosis. Furthermore, we explore its association with motor and nonmotor features and its value for phenoconversion and disease progression prediction.

**Methods:** PPMI data were downloaded for de novo PD subjects, subjects at-risk for developing a synucleinopathy (with isolated REM sleep behavior disorder, hyposmia or a pathogenic mutation) and controls. Impaired bed mobility was assessed with the MDS-UPDRS part 2 item 9. A frequency analysis was performed. Multivariable logistic regression analyses were used to investigate the association with other PD variables. Cox proportional-hazards models were used to test if difficulties with turning in bed could predict phenoconversion. Linear mixed models were used to evaluate if difficulties with turning in bed could predict disease progression.

**Results:** Of the at-risk subjects, 9.2-12.5% experienced difficulties with turning in bed vs. 25.0% of de novo PD subjects and 2.5% of controls. Impaired turning ability was associated with MDS-UPDRS motorscore (axial signs in the at-risk group, bradykinesia in the de novo PD group) and SCOPA-AUT score (gastrointestinal symptoms). In addition, difficulties with turning in bed were a significant predictor for phenoconversion in the at-risk group and for development of motor complications in the de novo PD group.

**Conclusion:** Our findings suggest that difficulties with turning in bed can be helpful as clinical symptom for a prodromal PD screening and for motor complication prediction in early PD.

**Highlights:**

- Subjective difficulties with turning in bed are a prodromal PD symptom.
- Impaired bed mobility predicts synucleinopathy phenoconversion in at-risk groups.
- Impaired bed mobility predicts development of motor complications in de novo PD.

## 1. Introduction

Sleep disturbances are disabling features of Parkinson’s disease (PD) and related disorders. Some of these symptoms, such as REM sleep behavior and excessive daytime sleepiness, are regarded as early or prodromal symptoms [1]. In contrast, nocturnal impaired bed mobility (impaired turning ability and supine body position during sleep) is considered to be a symptom of advanced disease [2]. However, recent studies using wearables or polysomnographies already show disturbances of body position and turning speed/ability in early and prediagnostic PD compared to control subjects [3,4]. These results suggest that impaired bed mobility is a very early PD sign. This raises the question if these differences are only detectable using wearables or if patients also experience impaired bed mobility in the prodromal phase. If so, this symptom might contribute to phenoconversion prediction in populations at-risk. Furthermore, impaired bed mobility is regarded as a nocturnal motor symptom and “off” phenomenon in advanced PD [2]. However, the association with other motor and nonmotor symptoms in early PD and its possible impact on disease progression is less investigated.

In this study, we investigate the frequency of subjective difficulties with turning in bed in patients with *de novo* PD and patients at-risk for developing a synucleinopathy. Furthermore, we investigate the factors associated with turning in bed in both groups. Finally, we investigate if impaired bed mobility predicts phenoconversion to a synucleinopathy in the at-risk group and if impaired bed mobility influences disease progression in the de novo PD group.

## 2. Methods

### 2.1 Population

The PPMI is an ongoing international multicenter observational cohort study investigating disease progression in PD with several PD populations and populations at-risk for developing PD and related disorders [5,6]. The methodology and details of the study assessments are available on the PPMI website (ppmi-info.org). The Institutional Review Board approved the PPMI program of each participating site and all participants gave their written informed consent to participate in the program.

For our study, we included three study groups:

1. Subjects with de novo PD. Inclusion criteria were a clinical diagnosis of PD [7] (resting tremor and/or bradykinesia and rigidity; or an asymmetric resting tremor/asymmetric bradykinesia), an age of ≥ 30 years at the time of PD diagnosis, time since diagnosis < 2 years, Hoehn and Yahr (H&Y) stage 1 or 2 [8], not expecting to require medication ≤ 6 months from baseline and a dopamine transporter deficit on neuroimaging. Exclusion criteria were the use of PD medication ≤ 60 days prior to baseline.
2. Unaffected subjects at-risk for developing a synucleinopathy with either: -a prodromal PD feature [9]: either a polysomnographic confirmed isolated REM sleep behavior disorder (RBD) or a University of Pennsylvania Smelling Identification Test (UPSIT) confirmed hyposmia [10]. Inclusion criteria were an age ≥ 60 years. Exclusion criteria were a clinically ni significant neurological or psychiatric disorder (including a clinical diagnosis of PD or dementia), a Geriatric Depression Scale (GDS) score ≥ 10 [11], a State-Trait Anxiety Inventory (STAI) score ≥ 54 [12] at baseline and a previously obtained MRI scan with evidence of a clinically significant neurological disorder. -a pathogenic mutation [*LRRK2, GBA* or *SNCA*]. Inclusion criteria were age ≥ years 45 for *LRRK2* and *GBA* and ≥ 30 years for *SNCA* mutation carriers. Exclusion criteria were a clinical PD diagnosis.
3. Control subjects. Inclusion criteria were ≥ 30 years at screening. Exclusion criteria were a clinically significant neurological disorder, a first-degree relative with idiopathic PD and a Montreal Cognitive Assessment (MoCA) score ≤ 26 [13].

Exclusion criteria for all groups were use of medication that could interfere with dopamine transporter imaging ≤ 6 months of screening, use of medication or medical conditions that were contra-indicated for a lumbar puncture, usage of investigational drugs or devices ≤ 60 days prior to baseline and development of an alternative diagnosis during follow-up.

### 2.2 Clinical assessment

The following demographics were evaluated: age, gender and time since diagnosis. Disease stage and severity were assessed with the H&Y stage and total scores of the Movement Disorder Society Unified Parkinson’s Disease rating scale (MDS-UPDRS) part 1 (nonmotor aspect daily living), 2 (motor aspect daily living) and 3 (motor examination) [14]. The nonmotor assessment included total scores of MoCA (cognition) [13], Scales for outcomes in Parkinson’s disease - autonomic dysfunction (SCOPA-AUT) [15], STAI (anxiety) [12], Epworth Sleepiness Scale (ESS) [16], RBD screening questionnaire [17], UPSIT (smell) [10] and GDS (depression) [11]. For the MDS-UPDRS part 3 and SCOPA-AUT, subscores were calculated: MDS-UPDRS part 3 tremor (items 15-18), rigidity (item 3), bradykinesia (items 2, 4-9 and 14) and axial (items 1, 9-13). SCOPA-AUT gastrointestinal symptoms (items 1-7), urinary dysfunction (items 8-13), cardiovascular dysfunction (items 14-16), thermoregulation (items 17-21) and sexual dysfunction (items 22-23 for males and 24-25 for females). Difficulties turning over in bed at baseline were assessed with the MDS-UPDRS part 2 item 9 and a score ≥ 1 was classified as positive. Subjects with missing data on this item were excluded. Follow-up visits were scheduled every 3 months in the first year and every 6 months afterwards. Data were downloaded in January 2021. We used the baseline data of all groups and the follow-up data for 5 years for the *de novo* PD group and follow-up data for 4 years for the at-risk group.

### 2.3 Statistical analyses

Frequency differences in bed-turning difficulties between groups at baseline were tested with a Chi-square test. Within the at-risk group and the de novo PD group, differences in motor and nonmotor variables scores were compared between subjects with and without bed-turning difficulties. Kolmogorov-Smirnov tests were used to test the distribution of all variables. In addition, t-tests were used for normally distributed continuous variables, Mann Whitney U-test for non-normally distributed continues variables and Chi-square tests for categorical variables.

To investigate which motor and nonmotor variables were associated with bed-turning difficulties, a multivariable logistic regression model with a forced entry design was used for the at-risk and de novo PD groups. Impaired turning ability in bed was used as dependent variable and all variables that showed a significant difference in mean/median scores between groups were used as independent variables. MDS-UPDRS part 1 and 2 scores were not included because of their overlap with other nonmotor scales and the dependant variable. For the independent variables that had a statistically significant association with bed-turning difficulties, post-hoc multivariable logistic regression analyses were performed to test which subscores were associated with bed-turning difficulties.

The at-risk group was followed up for 4 years and cox proportional-hazards models were used to test if impaired turning in bed at baseline could predict phenoconversion to a synucleinopathy (PD, dementia with Lewy bodies or Multiple System Atrophy). Phenoconversion was defined as an event when the diagnosis was consistent for ≥ 2 visits or occurred at the most recent visit. Time to event was defined as the period from baseline until event in years. In the absence of a conversion event, data were censored at the most recent visit. First, a univariable Cox proportional-hazard model with bed-turning difficulties at baseline as a variable was used. Secondly, we performed a multivariable Cox proportional-hazard model with also nonmotor variables (MOCA, SCOPA-AUT, STAI, ESS, RBD screening questionnaire, UPSIT and GDS) and age at baseline as covariates. Thirdly, a Cox proportional-hazard model with both motor variables (MDS-UPDRS part 3 total score) and nonmotor variables at baseline as covariates were used. Hazard ratios with 95% intervals were calculated and Kaplan-Meier estimates and curves were generated.

Linear mixed models (LMM) were used to test if impaired turning in bed at baseline could predict disease progression over 5 years in the de novo PD cohort. We fitted 3 restricted maximum likelihood LMMs with an autoregressive covariance structure and as dependent variables: 1. UPDRS part 3 total score, 2. MoCA and 3. UPDRS part 4 total score. To improve normality of distribution of the residuals, testing was based upon log-transformed values for the MoCA and UPDRS-4 scores. Difficulties with turning in bed and gender were included as fixed factors. Time, age, Levodopa equivalent daily dose (LEDD) and baseline scores of SCOPA-AUT, STAI, ESS, RBD screening questionnaire, UPSIT and GDS were added as fixed covariates. For the MDS-UPRDS-3 and MDS-UPDRS-4 model, the MoCA score at baseline was also added as a fixed covariate and for MoCA and UPDRS-4 models, the UPDRS-3 score at baseline was at baseline added as a fixed covariate. Finally, the interaction between time and difficulties with turning in bed and between time and LEDD were included as fixed factors. Random intercepts were used for each subject, and time and LEDD were also included as random factors.

A Bonferroni correction was used for multiple comparison correction, when appropriate. All analyses were performed with IBM SPSS Statistics for Mac (Version 26.0. Armonk, NY: IBM Corp.)

## 3. Results

### 3.1 Cross-sectional analyses

At baseline 421 subjects fulfilled the inclusion criteria in the de novo PD group vs 487 subjects in the at-risk group (64 subjects with a prodromal PD feature and 423 subjects with a pathogenic mutation) and 195 subjects in the control group (figure 1). The frequency analysis results are summarized in table 1. The de novo PD group had the highest frequency of difficulties with turning in bed (57.6%), followed by subjects with a prodromal PD feature (12.5%), subjects with a genetic mutation (9.2%) and controls (2.6%), p<0.001. Differences in clinical parameters between subjects with and without difficulties with turning in bed are summarized in table 2 for the at-risk group and table 3 for the de novo PD group.

**Table 1.** Frequency analysis results. describes the different frequencies of bed turning difficulties in each group.

|  | Total (N) | Bed turning difficulties | No bed turning difficulties | Percentage (95% confidence interval) |
| --- | --- | --- | --- | --- |
| Controls | 195 | 5 | 190 | 2.6% (CI 1.1%-5.9%) |
| Prodromal PD | 64 | 8 | 56 | 12.5% (CI 6.5%-22.8%) |
| Genetic unaffected PD | 423 | 39 | 384 | 9,2% (CI 6.8%-12.4%) |
| De novo PD | 421 | 108 | 313 | 25.6% (21.7-30.0%) |
| De novo PD after 5 years follow up | 314 | 181 | 133 | 57.6% (52.1-63.0%) |

PD=Parkinson's disease, N=number of subjects, CI= confidence interval. Differences between groups were tested with a Chi-square test: $p < 0.001$ .

**Table 2.** Clinical characteristics of the at-risk group. describes the baseline clinical characteristics of the at-risk group with and without bed turning difficulties. F=female, M=male, MDS-UPDRS=Movement Disorder Society-Unified Parkinson Disease Rating Scale, MoCA= Montreal Cognitive Assessment, RBD=REM sleep behavior disorder, UPSIT=University of Pennsylvania Smell Identification Test, SCOPA-AUT=Scales for outcome in Parkinson’s Disease-Autonomic dysfunction. STAI=State-Trait Anxiety Inventory, QUIP=questionnaire for Impulsive-Compulsive Disorder, ESS=Epworth Sleepiness scale. Mann Whitney U tests were used to test group differences for non-normally distributed continues variables and Chi-square tests* for categorial variables. A Bonferroni correction for multiple comparison was used (0.05/14=0.003) and significant P values are shown in bold.

| Variable | No turning difficulties<br>N= 440 | Turning difficulties<br>N= 47 | P value |
| --- | --- | --- | --- |
| Age (years) | 62 (56-67) | 66 (61-72) | **0.001** |
| Gender (F/M)* | 241/199 | 26/21 | 0.534 |
| Hoehn and Yahr | 0 (0-0) | 0 (0-0) | **0.001** |
| MDS-UPDRS 1 | 4 (2-7) | 10 (4-12) | **<0.001** |
| MDS-UPDRS 2 | 0 (0-1) | 4 (1-8) | **<0.001** |
| MDS-UPDRS 3 | 1 (0-4) | 3.5 (1-9) | **<0.001** |
| MoCA | 27 (25-29) | 26 (24-28) | 0.137 |
| RBD screening | 3 (1-5) | 4 (3-7) | **0.002** |
| UPSIT | 33 (29-36) | 33 (26-36) | 0.625 |
| SCOPA-aut | 7 (4-11) | 11 (6-20) | **<0.001** |
| Geriatric Depression | 1 (0-3) | 2 (1-4) | **0.001** |
| STAI | 58 (48-70) | 64 (46-80) | 0.196 |
| QUIP | 0 (0-1) | 0 (0-1) | 0.070 |
| ESS | 5 (3-7) | 5 (3-9) | 0.271 |

**Table 3.** Clinical characteristics of the de novo PD group. describes the baseline clinical characteristics of the novo Parkinson’s disease (PD) group with and without bed turning difficulties. F=female, M=male, MDS-UPDRS=Movement Disorder Society-Unified Parkinson Disease Rating Scale, MoCA= Montreal Cognitive Assessment, RBD=REM sleep behavior disorder, UPSIT=University of Pennsylvania Smell Identification Test, SCOPA-AUT=Scales for outcome in Parkinson’s Disease-Autonomic dysfunction STAI=State-Trait Anxiety Inventory, QUIP=questionnaire for Impulsive-Compulsive Disorder, ESS=Epworth Sleepiness scale. Mann Whitney U tests were used to test group differences for non-normally distributed continues variables and Chi-square tests* for categorial variables. A Bonferroni correction for multiple comparison was used (0.05/15=0.003) and significant P values are shown in bold.

| Variable | No turning difficulties<br>N= 313 | Turning difficulties<br>N=108 | P value |
| --- | --- | --- | --- |
| Age (years) | 62 (54-69) | 63 (55-68) | 0.384 |
| Gender (F/M)* | 101/212 | 43/65 | 0.154 |
| Hoehn and Yarh | 2 (1-2) | 2 (1-2) | 0.005 |
| UPDRS 1 | 4 (2-6) | 7 (5-10) | **<0.001** |
| UPDRS 2 | 4 (2-6) | 9 (6-13) | **<0.001** |
| UPDRS 3 | 18 (14-24) | 24 (18-30) | **<0.001** |
| MOCA | 28 (26-29) | 27 (26-29) | 0.461 |
| RBD screening | 3 (2-5) | 5 (3-7) | **<0.001** |
| UPSIT | 22 (17-28) | 23 (14-30) | 0.958 |
| SCOPA-aut | 8 (5-11) | 11 (8-15) | **<0.001** |
| Geriatric Depression | 1 (0-3) | 2 (1-3) | **<0.001** |
| STAI | 61 (50-75) | 67 (52-80) | 0.017 |
| QUIP | 0 (0-0) | 0 (0-0) | 0.947 |
| ESS | 5 (3-8) | 6 (3-9) | 0.063 |
| Time since diagnosis<br>(months) | 4 (2-8) | 4 (2-9) | 0.534 |

**Figure 1:**
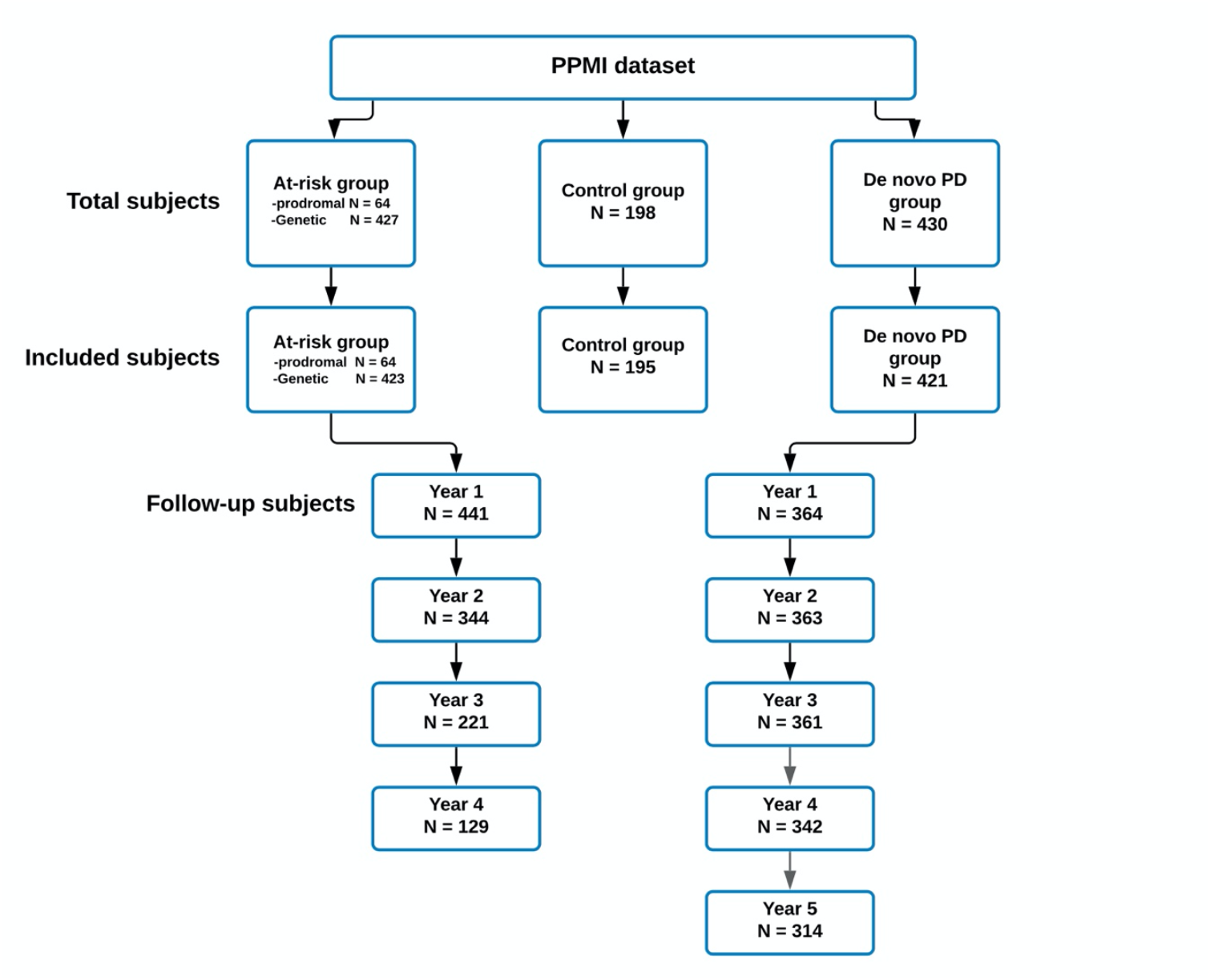
Overview of included PPMI subjects. Flowchart of included subjects. PPMI=Parkinson Progression Marker Initiative, PD=Parkinson’s disease, N=number.

The logistic regression analyses results are summarized in table 4. There was a significant effect of MDS-UPRDS-3 score (OR 1.1, p<0.001 de novo PD group; OR 1.1, p=0.001 at-risk group) and SCOPA-AUT score (OR 1.1, p=0.003 de novo PD group; OR 1.1, p=0.003 at-risk group) on difficulties with turning in bed in both groups. Using MDS-UPDRS 3 subscores, the effect was explained by axial symptoms in the at-risk group (OR=1.7, p<0.001) and by bradykinesia in the de novo PD group (OR 1.1, p=0.005). Using SCOPA-AUT subscores, the effect was explained by gastrointestinal symptoms in the de novo PD group (OR=1.2, p=0.001) with a trend towards significance in the at-risk group (OR 1.2, p=0.025 uncorrected).

**Table 4.** Predictors of bed turning difficulties. describes the different clinical variables that predict bed turning difficulties in the de novo Parkinson’s disease (PD) group and the at-risk group. MDS-UPDRS=Movement Disorder Society-Unified Parkinson Disease Rating Scale, RBD=REM sleep behavior disorder, SCOPA-AUT=Scales for outcome in Parkinson’s Disease-Autonomic dysfunction, STAI=State-Trait Anxiety Inventory. Multivariable logistic regression analyses were used with a Bonferroni correction for multiple comparisons (main analysis: 0.05/5=0.010, MDS-UPDRS-3 subscore analysis: 0.05/4=0.013, SCOPA-aut subscore analysis: 0.05/5=0.010). Significant P values are shown in bold.

| variable | De novo PD group |  | At-risk group |  |
| --- | --- | --- | --- | --- |
|  | OR (95% CI) | P | OR (95% CI) | P |
| MDS-UPDRS 3 | 1.1 (1.0-1.1) | **<0.001** | 1.1 (1.1-1.2) | **0.001** |
| RBD screening questionnaire | 1.1 (1.0-1.2) | 0.012 | 1.1 (1.0-1.2) | 0.354 |
| Geriatric Depression scale | 1.1 (1.0-1.2) | 0.211 | 1.1 (1.0-1.3) | 0.392 |
| SCOPA-AUT | 1.1 (1.0-1.1) | **0.003** | 1.1 (1.0-1.1) | **0.003** |
| STAI | 1.0 (1.0-1.0) | 0.848 | 1.0 (1.0-1.0) | 0.467 |
| **Post-hoc subscore analyses** |  |  |  |  |
| *MDS-UPDRS 3* |  |  |  |  |
| Tremor | 1.0 (0.9-1.0) | 0.323 | 1.1 (1.0-1.3) | 0.333 |
| Rigidity | 1.1 (1.0-1.2) | 0.042 | 1.0 (0.7-1.5) | 0.936 |
| Bradykinesia | 1.1 (1.0-1.1) | **0.005** | 1.1 (1.0-1.2) | 0.195 |
| Axial signs | 1,1 (1.0-1.3) | 0.076 | 1.7 (1.3-2.3) | **<0.001** |
| *SCOPA-AUT* |  |  |  |  |
| Urinary | 1.1 (1.0-1.2) | 0.079 | 1.0 (0.9-1.2) | 0.426 |
| Cardiovascular | 1.3 (1.0-1.8) | 0.085 | 1.3 (0.9-1.8) | 0.155 |
| Sexual | 1.0 (0.8-1.1) | 0.577 | 1.2 (1.0-1.4) | 0.118 |
| Gastrointestinal | 1.2 (1.1-1.5) | **0.001** | 1.2 (1.0-1.3) | 0.025 |
| Thermoregulatory | 1.0 (0.9-1.2) | 0.994 | 1.0 (0.9-1.2) | 0.500 |

### 3.2 Longitudinal analyses

In the at-risk group, cox proportional-hazard models were used to investigate if turning difficulties at baseline could predict phenoconversion to a synucleinopathy within 4 years. 71 subjects converted. Using a univariable model, difficulties with turning bed were a significant predictor of phenoconversion (HR 2.40, CI 1.28-4.45, p=0.006, figure 2). The effect remained significant when controlling for nonmotor variables, age and gender (HR 2.40, CI 1.20-4.78, p=0.013). However, when also correcting for UPDRS motor score, the statistically significant effect disappeared (HR 1.43, CI 0.67-3.05, p=0.358), indicating that difficulties with turning in bed are a predictor of phenoconversion independent from nonmotor symptoms but dependent on motor signs during the day.

**Figure 2:**
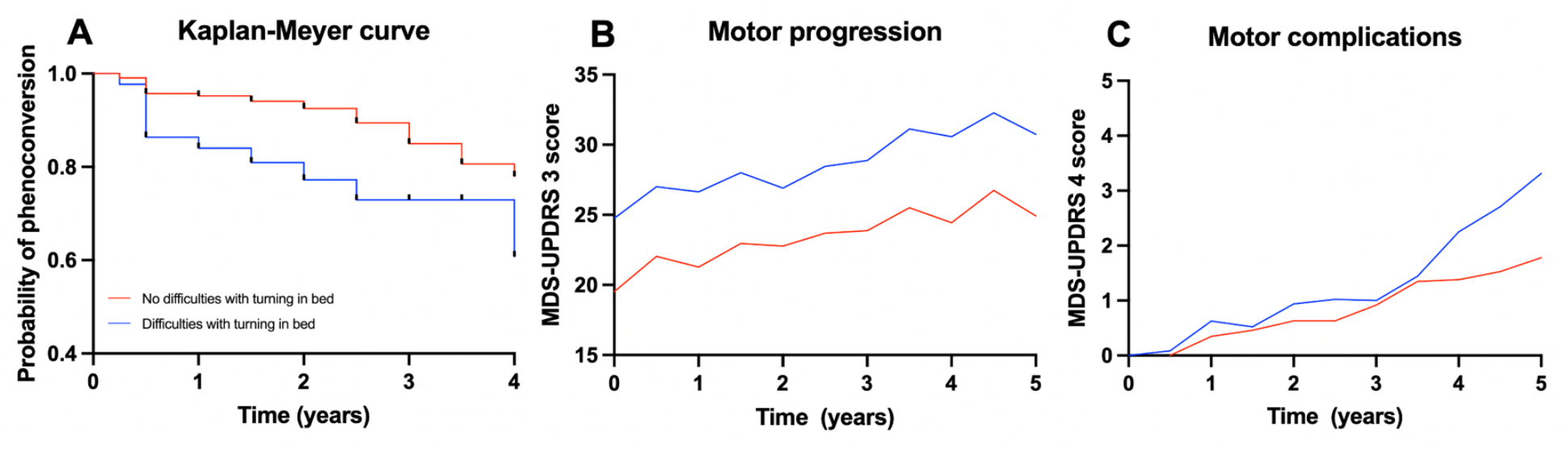
Longitudinal analyses results. Differences in disease course between subjects with and subjects without difficulties with turning in bed. A. is a Kaplan-Meyer curve that shows the probability of phenoconversion to a synucleinopathy in the at-risk group. B. shows the mean results of motor progression in the de novo PD group. C. shows the mean results of the development of motor complications in the de novo PD group. PD=Parkinson’s Disease. MDS-UPDRS=Movement Disorder Society-Unified Parkinson’s Disease Rating scale.

In the de novo PD group, LMMs were used to test if impaired turning in bed at baseline could predict disease progression over 5 years. The results are summarized in table 5 and figure 2. There was a significant effect of difficulties with turning in bed on UPDRS-3 score (ß 4.40, p<0.001) without a significant effect of the interaction between difficulties with turning in bed and time (ß -0.01, p=0.939): indicating that subjects with difficulties with turning in bed at baseline have more severe motor signs (at baseline and during follow up) without faster motor progression. There was no significant effect of difficulties with turning in bed or the interaction with time on the MoCA score (ß 0.02, p=0.139 and ß 0.00, p=0.103). There was no significant effect of difficulties with turning in bed on UPDRS-4 score (ß -0.21, p 0.015) with a statistically significant interaction between difficulties with turning in bed and time (ß 0.05, p<0.001): indicating that subjects with difficulties with turning in bed at baseline develop more severe motor complications within 5 years.

**Table 5.** The influence of bed turning difficulties on disease progression. describes the linear mixed model results of the de novo Parkinon’s disease (PD) group. MoCA= Montreal Cognitive Assessment. MDS-UPDRS=Movement Disorder Society-Unified Parkinson Disease Rating Scale. The analyses describe the effect of time, bed turning difficulties and their interaction on disease progression (motor progression [MDS-UPDRS-3], cognitive deterioration [MoCA] and motor complications [MDS-UPDRS-4]). The analyses were adjusted for gender, age, Levodopa Equivalent Daily Dose, baseline scores of Scales for outcome in Parkinson’s Disease-Autonomic dysfunction, REM sleep behavior disorder screening questionnaire, University of Pennsylvania Smell Identification Test, State-Trait Anxiety Inventory, Epworth Sleepiness scale and Geriatric Depression scale, the interaction between time and LEDD, MDS-UPRDS 3 (for MDS-UPDRS 4 and MoCA) and MoCA (for MDS-UPDRS 3 and 4). A Bonferroni correction was used for multiple comparison (0.05/9=0.006) and significant P values are shown in bold.

| Variable | $\beta$ (SE) | CI | P value |
| --- | --- | --- | --- |
| *MDS-UPDRS 3* |  |  |  |
| Time | 0.83 (0.17) | 0.49 - 1.18 | **<0.001** |
| Bed turning difficulties | 4.40 (1.03) | 2.37 - 6.43 | **<0.001** |
| Time x Bed turning difficulties | - 0.01 (0.18) | - 0.34 - 3.62 | 0.939 |
| *MoCA* |  |  |  |
| Time | - 0.01 (0.00) | - 0.00 - 0.01 | 0.020 |
| Bed turning difficulties | 0.02 (0.01) | - 0.01 - 0.04 | 0.139 |
| Time x Bed turning difficulties | 0.00 (0.00) | - 0.01 - 0.00 | 0.103 |
| *MDS-UPDRS 4* |  |  |  |
| Time | 0.08 (0.01) | 0.05 - 0.11 | **<0.001** |
| Bed turning difficulties | - 0.21 (0.08) | - 0.04 - 0.04 | 0.015 |
| Time x Bed turning difficulties | 0.05 (0.01) | 0.03 - 0.07 | **<0.001** |

## 4. Discussion

In this study, we investigated the frequency, associated symptoms, and predictive value of subjective difficulties with turning in bed in patients with the novo PD and patients at-risk to develop a synucleinopathy. Our main findings were a higher frequency of difficulties with turning in bed in both groups (25% and 9.2-12.5% vs 2.5% in the control group). In addition, difficulties with turning in bed were a predictor of phenoconversion in the at-risk group and a predictor of development of motor complications in the de novo PD group. Furthermore, difficulties with turning in bed were associated with motor signs (during the day) and autonomic symptoms. These findings combined suggest that difficulties with turning in bed are a very early PD symptom, that is easy to ask to patients and may be useful in clinical practice as part of prodromal PD screening and in disease progression prediction.

### 4.1 Associated symptoms

Our results show an association between difficulties with turning in bed and motor signs (evaluated by MDS-UPDRS 3) and autonomic dysfunction (assessed by SCOPA-AUT). These findings are in line with Mirelman et al. [3], who described an association between reduced nocturnal movements and turning velocity (evaluated by sensors) in early PD. Our results additionally show that this association also relates to subjective turning difficulties and is already present before PD diagnosis.

The association with motor signs during the day confirms that turning bed difficulties are an early nocturnal motor symptom and both are correlated. Using motor subscores, difficulties with turning in bed were mainly associated with axial signs in the at-risk group (OR 1.7, P<0.001). This seems logical considering the role of axial movements in turning in bed and is in line with the previously described correlation between impaired bed mobility and axial signs in PD [2,18]. Difficulties with turning in bed in the de novo group were mainly associated with bradykinesia (OR 1.1, p=0.005), suggesting nocturnal bradykinesia as etiology. This might also be explained by the higher frequency of bradykinesia in the de novo PD group, since bradykinesia is an essential criterion for PD diagnosis.

The relation between impaired bed mobility and autonomic dysfunction is not clear. Previous studies showed a negative influence of autonomic dysfunction on sleep disturbances [19] and an association with RBD [20]. This influence might also involve sleep-related and nocturnal movements. When using autonomic subscores, difficulties with turning in bed were mainly associated with gastrointestinal dysfunction (p=0.001 in the de novo group and p=0.025 [uncorrected] in the at-risk group). This might suggest a pathophysiological involvement of the gut-brain axis [21].

### 4.2 Disease progression

Our results show that patients that are at-risk of developing a synucleinopathy and who experience difficulties with turning in bed, more often phenoconvert than patients at-risk without difficulties with turning in bed within 4 years (HR 2.4, P=0.006). These findings are in line with Fereshtehnejad et al., who showed that patients with isolated RBD who phenoconvert, start to develop difficulties with turning in bed around 4 years prior to PD diagnosis compared to healthy control subjects without RBD [22]. Our findings additionally show that this is also the case for other (genetic) at-risk groups and suggest that difficulties with turning in bed might be useful as a phenoconversion predictor within at-risk groups. When controlling for other features, difficulties with turning in bed were a significant predictor independently of nonmotor features. However, when controlling for MDS-UPDRS-3 score, the significant effect disappeared: suggesting that motor signs during the day modulate the predictive effect. This may be explained by the association between both as described above. This finding highlights the clinical relevance of evaluating extrapyramidal signs, especially axial signs, in patients who complain about difficulties with turning in bed in sleep medicine.

Within the de novo PD group, patients who experienced difficulties with turning in bed developed more severe motor complications than patients without difficulties with turning in bed within 5 years (ß =0.05, p<0.001), independent of other motor and nonmotor variables and LEDD. Previous studies described that several factors can predict development of motor complications in PD: medication factors (LEDD and therapy response), neuropsychiatric factors (apathy, mood changes and anxiety), motor factors (MDS-UPDRS-2 total score) and patients related factors (age at onset, BMI, education status) [23,24]. Our study adds difficulties with turning in bed as an independent predictor. This might increase specificity in identifying which patients are at-risk of developing motor complications, which could have therapeutic consequences in clinical practice.

The relation between difficulties with turning in bed and the development of motor complications is unclear. In a cross-sectional study, Schaeffer et al. reported that MDS-UPDRS 4 score predicts subjective nocturnal immobility [25]. Our longitudinal results, however, suggest an inverse relation. This could be a causal relation, in which impaired bed mobility reduces sleep quality, that might influence development of motor complications. However, until now no other sleep-related symptoms have been associated with development of motor complications. Alternatively, patients that experience difficulties with turning in bed, might be more aware or focused on their motor symptoms and might notice motor complications earlier, since both MDS-UPDRS 2 and 4 are subjective scales [25]. Future studies that use objective outcome measures (such as sensors) might give more insight in this relation.

### 4.3 Limitations

Our study has several limitations. First, our study aimed to investigate subjective nocturnal immobility, in which nocturnal immobility was evaluated using the MDS-UPDRS-2 questionnaire. Although our results clearly show that a significant amount of patients are aware of this symptom in both groups, the frequency is lower than in studies that used objective measures such as sensors and polysomnographies [3,4]. Consequently, our results may be an underestimation of true nocturnal bed immobility in (prodromal) PD. Usage of additional sensors would probably increase sensitivity and specificity and correct for misperception in nocturnal bed immobility evaluation.

Furthermore, the relative high number of missing follow-up data in the at-risk group could have caused a selection bias for the survival analysis. The relatively low sample size of LBD and MSA subjects also did not allow us to discriminate between the different synucleinopathies. Finally, the follow-up data are limited to 4 and 5 years, so no conclusions can be drawn about long-term prediction.

## 5. Conclusion and further directions

In conclusion, our study confirms that subjective impaired bed mobility is an early PD symptom, that predicts phenoconversion in patients at-risk to develop a synucleinopathy and predicts the development of motor complications in patients with de novo PD. Future studies should evaluate the additional value of subjective and objective impaired bed mobility in the general population as prodromal PD signs and explore the association between impaired bed mobility and development of motor complications.

## Data Availability

The dataset is available at https://www.ppmi-info.org/

## 10. Acknowledgements

We like to thank all patients and healthy volunteers for participating in the PPMI study, all investigators who contributed, and the Michael J. Fox Foundation. For up-to-date information on the survey, visit http://www.ppmi-info.org/data. Furthermore, we would like to thank Kristien Wouters (medical statistics, Antwerp University Hospital).

## 11. Author roles

Conception and design of the study: *FD, IdV, MV, PC, DC*, Data analysis: *FD*, drafting the article: *FD*, revising the article for important intellectual content: *IdV, MV, PC, DC*.

## References

[1] A. Al-Qassabi, S.-M. Fereshtehnejad, R.B. Postuma, Sleep Disturbances in the Prodromal Stage of Parkinson Disease, Curr Treat Options Neurol. 19 (2017) 22. https://doi.org/10.1007/s11940-017-0458-1.

[2] R. Bhidayasiri, C. Trenkwalder, Getting a good night sleep? The importance of recognizing and treating nocturnal hypokinesia in Parkinson’s disease, Parkinsonism Relat Disord. 50 (2018) 10–18. https://doi.org/10.1016/j.parkreldis.2018.01.008.

[3] A. Mirelman, I. Hillel, L. Rochester, S. Del Din, B.R. Bloem, L. Avanzino, A. Nieuwboer, I. Maidan, T. Herman, A. Thaler, T. Gurevich, M. Kestenbaum, A. Orr-Urtreger, M. Brys, J.M. Cedarbaum, N. Giladi, J.M. Hausdorff, Tossing and Turning in Bed: Nocturnal Movements in Parkinson’s Disease, Movement Disorders. 35 (2020) 959–968. https://doi.org/10.1002/mds.28006.

[4] F. Dijkstra, N. Reyn, B. de Bruyn, K. van den Bossche, I. de Volder, M. Willemen, M. Viaene, null Jo Leenders, P. Cras, D. Crosiers, REM sleep without atonia and nocturnal body position in prediagnostic Parkinson’s disease, Sleep Med. 84 (2021) 308–316. https://doi.org/10.1016/j.sleep.2021.06.011.

[5] K. Marek, D. Jennings, S. Lasch, A. Siderowf, C. Tanner, T. Simuni, C. Coffey, K. Kieburtz, E. Flagg, S. Chowdhury, W. Poewe, B. Mollenhauer, P.-E. Klinik, T. Sherer, M. Frasier, C. Meunier, A. Rudolph, C. Casaceli, J. Seibyl, S. Mendick, N. Schuff, Y. Zhang, A. Toga, K. Crawford, A. Ansbach, P. De Blasio, M. Piovella, J. Trojanowski, L. Shaw, A. Singleton, K. Hawkins, J. Eberling, D. Brooks, D. Russell, L. Leary, S. Factor, B. Sommerfeld, P. Hogarth, E. Pighetti, K. Williams, D. Standaert, S. Guthrie, R. Hauser, H. Delgado, J. Jankovic, C. Hunter, M. Stern, B. Tran, J. Leverenz, M. Baca, S. Frank, C.-A. Thomas, I. Richard, C. Deeley, L. Rees, F. Sprenger, E. Lang, H. Shill, S. Obradov, H. Fernandez, A. Winters, D. Berg, K. Gauss, D. Galasko, D. Fontaine, Z. Mari, M. Gerstenhaber, D. Brooks, S. Malloy, P. Barone, K. Longo, T. Comery, B. Ravina, I. Grachev, K. Gallagher, M. Collins, K.L. Widnell, S. Ostrowizki, P. Fontoura, T. Ho, J. Luthman, M. van der Brug, A.D. Reith, P. Taylor, The Parkinson Progression Marker Initiative (PPMI), Progress in Neurobiology. 95 (2011) 629–635. https://doi.org/10.1016/j.pneurobio.2011.09.005.

[6] K. Marek, S. Chowdhury, A. Siderowf, S. Lasch, C.S. Coffey, C. Caspell-Garcia, T. Simuni, D. Jennings, C.M. Tanner, J.Q. Trojanowski, L.M. Shaw, J. Seibyl, N. Schuff, A. Singleton, K. Kieburtz, A.W. Toga, B. Mollenhauer, D. Galasko, L.M. Chahine, D. Weintraub, T. Foroud, D. Tosun-Turgut, K. Poston, V. Arnedo, M. Frasier, T. Sherer, The Parkinson’s progression markers initiative (PPMI) – establishing a PD biomarker cohort, Annals of Clin. and Trans. Neur. 5 (2018) 1460–1477. https://doi.org/10.1002/acn3.644.

[7] D. Berg, C.H. Adler, B.R. Bloem, P. Chan, T. Gasser, C.G. Goetz, G. Halliday, A.E. Lang, S. Lewis, Y. Li, I. Liepelt-Scarfone, I. Litvan, K. Marek, C. Maetzler, T. Mi, J. Obeso, W. Oertel, C.W. Olanow, W. Poewe, S. Rios-Romenets, E. Schäffer, K. Seppi, B. Heim, E. Slow, M. Stern, I.O. Bledsoe, G. Deuschl, R.B. Postuma, Movement disorder society criteria for clinically established early Parkinson’s disease, Movement Disorders. 33 (2018) 1643–1646. https://doi.org/10.1002/mds.27431.

[8] M.M. Hoehn, M.D. Yahr, Parkinsonism: onset, progression and mortality, Neurology. 17 (1967) 427–442.

[9] D. Berg, R.B. Postuma, C.H. Adler, B.R. Bloem, P. Chan, B. Dubois, T. Gasser, C.G. Goetz, G. Halliday, L. Joseph, A.E. Lang, I. Liepelt-Scarfone, I. Litvan, K. Marek, J. Obeso, W. Oertel, C.W. Olanow, W. Poewe, M. Stern, G. Deuschl, MDS research criteria for prodromal Parkinson’s disease, Movement Disorders. 30 (2015) 1600–1611. https://doi.org/10.1002/mds.26431.

[10] R. Doty, The Smell Identification Test, TM administration manual, 3rd edn. Sensonics Inc., Philadephia (1995).

[11] J.A. Yesavage, T.L. Brink, T.L. Rose, O. Lum, V. Huang, M. Adey, V.O. Leirer, Development and validation of a geriatric depression screening scale: a preliminary report, J Psychiatr Res. 17 (1982) 37–49. https://doi.org/10.1016/0022-3956(82)90033-4.

[12] C. Spielberge, Manual for the State-trait Anxiety Inventory STAI (Form Y), Mind Garden; Palo Alto, CA: 1983. (n.d.).

[13] Z.S. Nasreddine, N.A. Phillips, V. Bédirian, S. Charbonneau, V. Whitehead, I. Collin, J.L. Cummings, H. Chertkow, The Montreal Cognitive Assessment, MoCA: A Brief Screening Tool For Mild Cognitive Impairment, Journal of the American Geriatrics Society. 53 (2005) 695– 699. https://doi.org/10.1111/j.1532-5415.2005.53221.x.

[14] C.G. Goetz, B.C. Tilley, S.R. Shaftman, G.T. Stebbins, S. Fahn, P. Martinez-Martin, W. Poewe, C. Sampaio, M.B. Stern, R. Dodel, B. Dubois, R. Holloway, J. Jankovic, J. Kulisevsky, A.E. Lang, A. Lees, S. Leurgans, P.A. LeWitt, D. Nyenhuis, C.W. Olanow, O. Rascol, A. Schrag, J.A. Teresi, V. Hilten, J.J N. LaPelle, Movement Disorder Society-sponsored revision of the Unified Parkinson’s Disease Rating Scale (MDS-UPDRS): Scale presentation and clinimetric testing results, Movement Disorders. 23 (2008) 2129–2170. https://doi.org/10.1002/mds.22340.

[15] M. Visser, J. Marinus, A.M. Stiggelbout, J.J. Van Hilten, Assessment of autonomic dysfunction in Parkinson’s disease: The SCOPA-AUT, Movement Disorders. 19 (2004) 1306– 1312. https://doi.org/10.1002/mds.20153.

[16] M.W. Johns, A new method for measuring daytime sleepiness: the Epworth sleepiness scale, Sleep. (1991) 540–545.

[17] K. Stiasny-Kolster, G. Mayer, S. Schäfer, J.C. Möller, M. Heinzel-Gutenbrunner, W.H. Oertel, The REM sleep behavior disorder screening questionnaire—A new diagnostic instrument, Movement Disorders. 22 (2007) 2386–2393. https://doi.org/10.1002/mds.21740.

[18] M.J. Steiger, P.D. Thompson, C.D. Marsden, Disordered axial movement in Parkinson’s disease, J Neurol Neurosurg Psychiatry. 61 (1996) 645–648. https://doi.org/10.1136/jnnp.61.6.645.

[19] J.A. Albers, P. Chand, A.M. Anch, Multifactorial sleep disturbance in Parkinson’s disease, Sleep Med. 35 (2017) 41–48. https://doi.org/10.1016/j.sleep.2017.03.026.

[20] R.B. Postuma, P.A. Lanfranchi, H. Blais, J.-F. Gagnon, J.Y. Montplaisir, Cardiac autonomic dysfunction in idiopathic REM sleep behavior disorder, Movement Disorders. 25 (2010) 2304–2310. https://doi.org/10.1002/mds.23347.

[21] H. Braak, R.A.I. de Vos, J. Bohl, K. Del Tredici, Gastric α-synuclein immunoreactive inclusions in Meissner’s and Auerbach’s plexuses in cases staged for Parkinson’s disease-related brain pathology, Neuroscience Letters. 396 (2006) 67–72. https://doi.org/10.1016/j.neulet.2005.11.012.

[22] S.-M. Fereshtehnejad, C. Yao, A. Pelletier, J.Y. Montplaisir, J.-F. Gagnon, R.B. Postuma, Evolution of prodromal Parkinson’s disease and dementia with Lewy bodies: a prospective study, Brain. 142 (2019) 2051–2067. https://doi.org/10.1093/brain/awz111.

[23] M.J. Kelly, M.A. Lawton, F. Baig, C. Ruffmann, T.R. Barber, C. Lo, J.C. Klein, Y. Ben-Shlomo, M.T. Hu, Predictors of motor complications in early Parkinson’s disease: A prospective cohort study, Movement Disorders. 34 (2019) 1174–1183. https://doi.org/10.1002/mds.27783.

[24] D. Santos-García, T. de Deus Fonticoba, E. Suárez Castro, A. Aneiros Díaz, D. McAfee, M.J. Catalán, F. Alonso-Frech, C. Villanueva, S. Jesús, P. Mir, M. Aguilar, P. Pastor, J. García Caldentey, E. Esltelrich Peyret, L.L. Planellas, M.J. Martí, N. Caballol, J. Hernández Vara, G. Martí Andrés, I. Cabo, M.A. Ávila Rivera, L. López Manzanares, N. Redondo, P. Martinez-Martin, COPPADIS Study Group, D. McAfee, Non-motor symptom burden is strongly correlated to motor complications in patients with Parkinson’s disease, Eur J Neurol. 27 (2020) 1210–1223. https://doi.org/10.1111/ene.14221.

[25] E. Schaeffer, T. Vaterrodt, L. Zaunbrecher, I. Liepelt-Scarfone, K. Emmert, B. Roeben, M. Elshehabi, C. Hansen, S. Becker, S. Nussbaum, J.-H. Busch, M. Synofzik, D. Berg, W. Maetzler, Effects of Levodopa on quality of sleep and nocturnal movements in Parkinson’s Disease, J Neurol. 268 (2021) 2506–2514. https://doi.org/10.1007/s00415-021-10419-7.

